# Organisational readiness, facilitators and barriers to the uptake of intravenous iron for the treatment of maternal anaemia in Nigeria: a mixed-methods pre-implementation assessment

**DOI:** 10.64898/2026.02.11.26346091

**Authors:** Mobolanle Balogun, Chisom Obi-Jeff, Yusuf Adelabu, Bosede Bukola Afolabi, Charles Ameh

**Affiliations:** Department of International Public Health, Liverpool School of Tropical Medicine, Liverpool, UK; Department of Community Health & Primary Care, Faculty of Clinical Sciences, College of Medicine, University of Lagos, Lagos, Nigeria; Department of Research, Brooks Insights Limited, Federal Capital Territory, Nigeria; Department of Medicine, Faculty of Clinical Sciences, College of Medicine, University of Lagos, Lagos, Nigeria; Department of Obstetrics & Gynaecology, Faculty of Clinical Sciences, College of Medicine, University of Lagos, Lagos, Nigeria; Department Obstetrics & Gynaecology, University of Nairobi, Nairobi, Kenya

## Abstract

**Background:** Maternal anaemia remains a leading cause of maternal morbidity and mortality in low- and middle-income countries (LMICs), with Nigeria bearing a disproportionate burden. Although treatment with intravenous (IV) iron formulations such as ferric carboxymaltose offer clinical advantages over oral iron, their uptake in routine maternal care in LMICs is limited.

**Objective:** This study critically assessed organisational readiness and identified context-specific facilitators and barriers to the adoption of IV iron therapy for maternal anaemia in Nigeria, using a mixed-methods pre-implementation design.

**Methods:** The study was conducted across six diverse healthcare facilities in Lagos, Nigeria. The validated Organisational Readiness for Implementing Change (ORIC) survey was administered to 74 healthcare providers in the facilities. Qualitative data were collected through 16 key informant interviews and 11 focus group discussions, guided by the Consolidated Framework for Implementation Research.

**Results:** High levels of organisational readiness were observed in the six facilities. Mean ORIC scores were 56.8 for pregnancy and 56.6 for postpartum (maximum score: 60), with no significant differences across facility types or healthcare cadres (p>0.05). Change commitment and change efficacy subscales showed consistently high scores (23.7/25 and 33/35 respectively). Key facilitators included perceived effectiveness of IV iron, relative advantage over other alternatives, compatibility with existing workflows and policies, and strong provider motivation. Primary barriers were cost concerns beyond the project period, risk of side effects, infrastructure and resource limitations, and low patient awareness. The facilitators identified in this study appeared to buffer against structural barriers, maintaining high readiness despite resource constraints.

**Conclusion:** Nigerian health facilities demonstrate substantial organisational readiness to adopt IV iron for maternal anaemia. Strategic implementation planning—addressing cost, infrastructure, and education will be critical to translating readiness into sustained practice. These findings offer transferable insights for scaling maternal health innovations in resource-constrained settings and contribute to global implementation science.

## Introduction

Anaemia is one of the most prevalent and preventable causes of morbidity and mortality during pregnancy and the post-partum period, affecting approximately 36% of pregnancies globally (1). The burden is higher in low- and middle-income countries (LMICs), with over 40% of pregnant women affected(1,2). Postpartum anaemia compounds this burden, with prevalence rates ranging from 50 to 80% across LMICs (3). In Nigeria, an estimated 45% of pregnant women experience moderate to severe anaemia (4). Iron deficiency anaemia (IDA) is the most common nutritional disorder in pregnancy and significantly affects maternal and fetal health (5,6).

Conventional management of IDA often involves oral iron supplementation; however, issues such as poor adherence, gastrointestinal side effects, and delays in haemoglobin correction hinder its effectiveness (7,8). Intravenous (IV) iron formulations offer several advantages, including rapid correction of anaemia, reduced frequency of healthcare visits, and elimination of adherence barriers associated with daily oral therapy (9). While IV iron is routinely used in high-income countries and supported by clinical practice guidelines(10,11), its adoption in LMICs like Nigeria remains limited. Contributing factors include healthcare system limitations, cost concerns, inadequate provider experience, and historical safety concerns associated with older IV iron formulations(12,13).

Recent hybrid trials have demonstrated that ferric carboxymaltose (FCM), a newer IV iron preparation, is effective, safe, and well-accepted for treating anaemia during pregnancy and postpartum in Nigeria (14–16). These favourable outcomes underscore FCM’s potential for broader adoption in real-world settings. However, critical questions remain regarding the readiness of health systems to implement IV iron in routine practice and the context-specific determinants influencing its uptake.

According to the Expert Recommendations for Implementing Change (ERIC) framework, assessing health system/organisational readiness, along with identification of barriers and facilitators, is a key implementation strategy to ensure successful adoption of evidence-based interventions (EBIs) (17). Despite this recommendation, organisational readiness assessments are infrequently conducted before the introduction of EBIs in LMICs. This gap is particularly pronounced for complex clinical interventions like IV iron that require specialised training, infrastructure modifications, and regular monitoring (18). The absence of readiness assessment increases the risk of suboptimal implementation or intervention failure (19). Organisational readiness for change is the shared resolve (change commitment) and collective ability (change efficacy) of organisational members to implement change (20). According to Weiner’s organisational readiness for change theory, factors influencing organisational readiness include change valence (members’ value of the change) and informational assessment (task demands, resource perception and situational factors) (19). Organisations with higher levels of readiness have more positive implementation outcomes and effective quality improvement implementation (21,22).

The Implementation Research for IV Iron Use in Pregnant and Postpartum Women in Nigeria project (IVON-IS) is currently examining FCM among pregnant and postpartum women, using continuous quality improvement (CQI) and participatory action research approaches to strengthen local anaemia care pathways (23). Before real-world implementation, robust assessment of organisational readiness and identification of system-level facilitators and barriers are needed to guide effective implementation strategy selection. Therefore, the objectives of this study were to evaluate organisational readiness and to identify key facilitators and barriers to adoption of IV iron for treating anaemia in pregnancy and the postpartum period within the Nigerian health system.

## Methods

### Study setting

This study was conducted in six healthcare facilities on the IVON-IS project in Lagos State, southwest Nigeria (23). The facilities all provide 24-hour antenatal, delivery and postpartum care to varying degrees. They were purposively selected as a cluster to represent the Nigerian healthcare system and its referral pattern. They comprised one comprehensive primary healthcare facility, two secondary healthcare facilities, one tertiary healthcare facility, and two private healthcare facilities. At the time this study was conducted, implementation management teams (IMTs) comprising different cadres of health care providers (HCPs) had been identified in each facility for the CQI process. The IMT and most of the other HCPs in maternal health units had been trained on FCM administration for moderate to severe anaemia by the IVON-IS team, either at a main training or step-down at the health facilities.

### Study design

The study used a mixed-methods evaluation where quantitative and qualitative data were collected to address study objectives. For the quantitative phase, the health facilities’ readiness to implement IV iron was determined by using the Organisational Readiness for Implementing Change (ORIC) survey, which is a 12-item instrument used to determine how well employees at an organisation feel they can implement the change in processes required by a proposed intervention (20). The instrument is divided into two facets- change commitment (5 items) and change efficacy (7 items) and is favoured for its brevity and basis on Weiner’s theory (19,20). Each item includes a Likert scale from 1 (Disagree) to 5 (Agree). The scale was used to assess readiness separately for FCM administration in pregnancy and in the postpartum.

For the qualitative phase, determinants (barriers and facilitators) of implementing routine use of IV iron for the treatment of anaemia during pregnancy and postpartum were evaluated using interviews guided by the 2009 version of the Consolidated Framework for Implementation Research (CFIR). CFIR is a determinant/explanatory framework comprising 39 constructs across five domains: intervention characteristics, outer setting, inner Setting, characteristics of individuals, and process (24).

### Study population and participant sampling

This study involved providers of antenatal and postnatal care at the six healthcare facilities. We excluded HCPs who were on leave or out of station during the study period.

Survey: A total population of 118 trained HCPs providing antenatal and postnatal care in the six facilities was targeted for the quantitative survey.

Key informant interviews (KIIs): We conducted KIIs with purposively targeted HCPs in leadership roles. At the PHC, we interviewed the medical officer of health, the apex nurse, the apex community health officer, and the senior medical officer. At the secondary level, we interviewed the heads of the obstetrics and gynaecology (O&G) departments and senior matrons in both health facilities, and the deputy managing director in one of the facilities. At the tertiary level, we interviewed the head of clinical services, the head of the O&G department and the senior matron. At the private hospitals, we interviewed the medical directors and senior matrons in both facilities. In total, 16 KIIs were conducted across the six facilities.

Focus group discussions (FGDs): All doctors and nurses who were expected to administer FCM directly were invited to participate in FDGs. The exception was the PHC where the FGD was only among nurses because it had just one other doctor apart from the senior medical officer. Eleven FGDs were conducted with the number of participants ranging from 4 to 8 and totaling 64 participants.

The number of KIIs (n=16) and FGDs (n=11) was determined using the information power concept (25).

### Data collection

Quantitative survey was self-administered and was collected electronically using REDCap, a secure web-based application hosted at the Centre for Clinical Trials, Research and Implementation Science, College of Medicine, University of Lagos (26,27). A link was generated to the survey tool that was sent to study participants along with consent forms.

The FGDs and KIIs were face-to-face and facilitated by MB or trained research assistants. Each interview lasted between 30 and 120 minutes and they were conducted in venues that were comfortable, quiet and private. All data was collected between August 14 and September 5, 2023.

### Data management and analysis

Quantitative data: Survey data was verified in real-time on REDCap by MB, research administrator and data manager. Completed data was extracted in Microsoft Excel and exported to STATA version SE 15.1 (StataCorp College Station, Texas, USA). Relevant summary and descriptive statistics were used for analysis. ORIC scores by cadre and facility type were compared using ANOVA.

Qualitative data: Data obtained from 27 audio recordings of interviews were transcribed verbatim and uploaded into ATLAS.ti 24.2.1 qualitative data analysis software. The codes were generated using an inductive approach based on Braun and Clarke’s thematic analysis approach (28). MB and COJ randomly selected three transcripts for content comprehension and familiarisation and independently coded them. The independent codes were compared and merged in a codebook that was used to code other transcripts line-by-line. Any new codes were discussed and included in the codebook until no further codes were generated. These codes were further reviewed and deductively grouped into the updated CFIR 2022 constructs as subthemes, and the domains as themes (29). The updated CFIR 2022 was released after our data collection, which was guided by the 2009 CFIR; thus, our open coding allowed us to adequately map subthemes and themes in the updated framework. Cross-analysis was done across the different levels of health care. We evaluated the quality of the results by applying the criteria of a framework on credibility, dependability, confirmability and transferability (30). (Table 1)

**Table 1:**
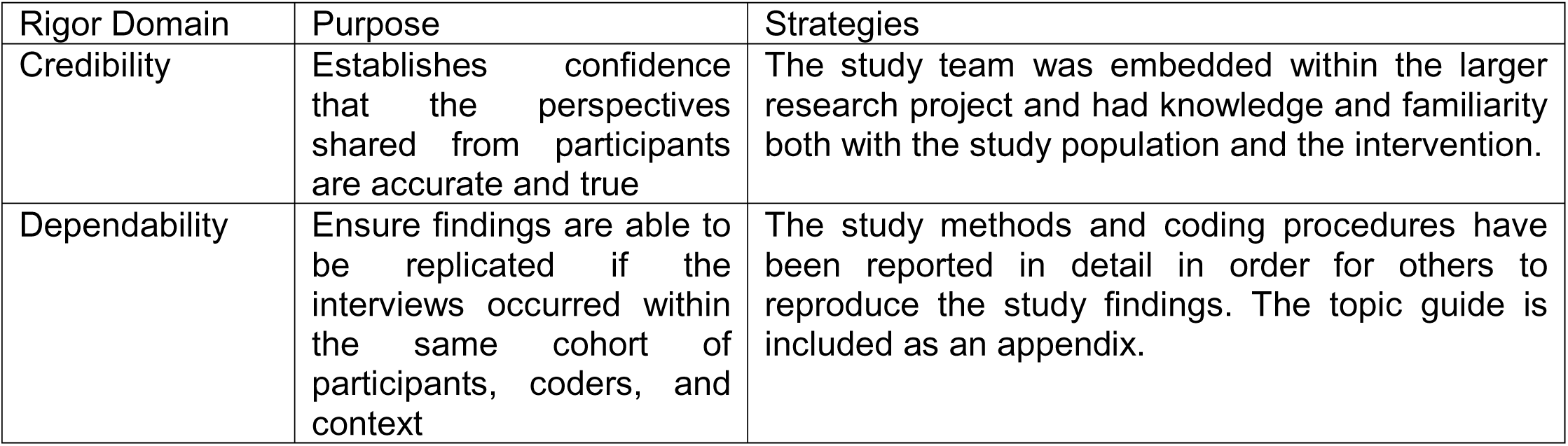
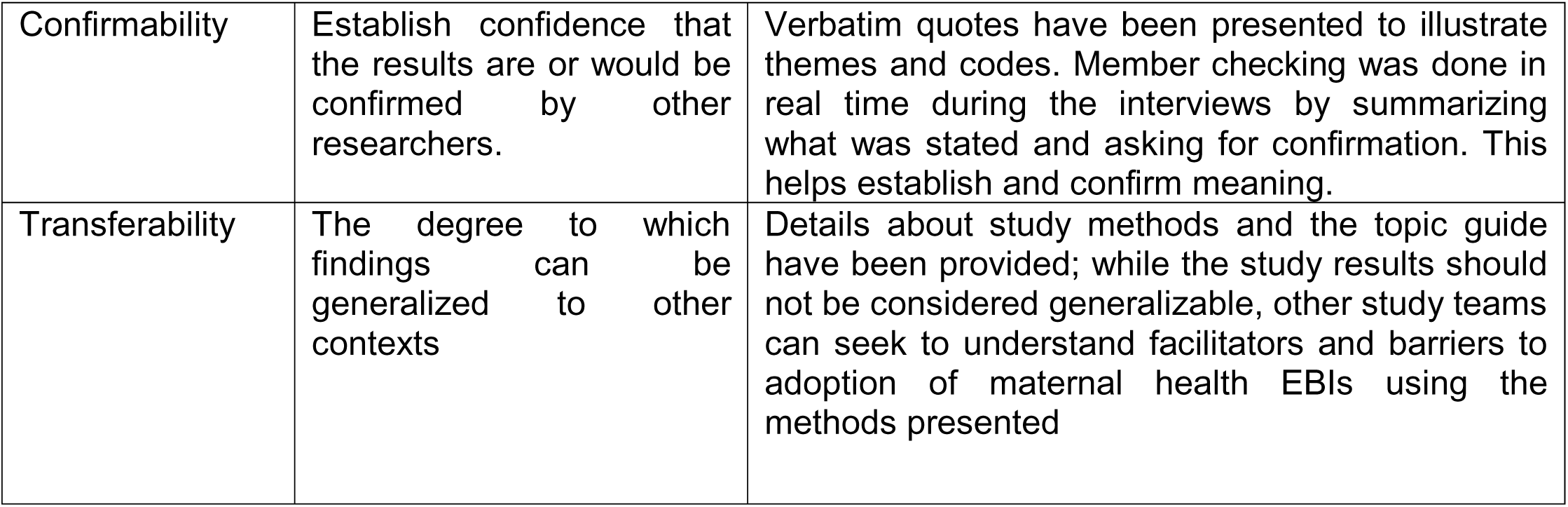
Strategies to enhance credibility and trustworthiness in the study.

### Ethical considerations

Ethical approval was obtained from the Health Research and Ethics Committee of the Lagos University Teaching Hospital (ADM/DCST/HREC/APP/5328) and Federal Medical Centre, Ebute Metta (HREC 22-22). Administrative approvals were obtained from the Lagos State Health Service Commission and the Lagos State Primary Health Care Board. Electronic consent was obtained for the surveys while verbal consent was obtained for the qualitative interviews. Confidentiality was maintained by not using identifiers.

## Results

### Quantitative results

#### Characteristics of respondents

Overall, 63% (74/118) of HCPs responded to the survey. The mean age of the respondents was 40.7 years (SD:10). Most of the HCPs are female (81.1%) and have completed tertiary education (75.7%). About an equal proportion (∼30%) were working in primary, secondary, and tertiary facilities and about half (51.4%) were nurses/midwives (Table 2).

**Table 2:** Personal and occupational characteristics of respondents.

| **Variables** | **Frequency (%)** |
| --- | --- |
| *Mean age in years (SD)* | 40.7 (10) |
| *Sex* |  |
| Female | 60 (81.1) |
| Male | 14 (18.9) |
| *Education* |  |
| Tertiary | 56 (75.7) |
| Postgraduate | 18 (24.3) |
| *Mean number of years in service (SD)* | 13.4 (9.3) |
| *Facility type* |  |
| Tertiary | 20 (27.0) |
| Secondary | 22 (29.7) |
| Primary | 23 (31.1) |
| Private | 9 (12.2) |
| *Job cadre* |  |
| Nurse/midwife | 38 (51.4) |
| Doctor | 10 (13.5) |
| Pharmacist | 8 (10.8) |
| Laboratory personnel | 6 (8.1) |
| Health information management personnel | 12 (16.2) |

### Organisational readiness for intravenous iron administration

The mean ORIC scores for IV iron use in pregnancy and postpartum were 56.8(SD:5.1) and 56.6(SD:4.9), respectively (range: 36-60, maximum score: 60). Scores for change commitment and change efficacy facets were equally high for IV iron use in pregnancy and postpartum (mean score%: 94.8% and 94.3%, respectively) [Table 3]. There were no statistically significant differences in ORIC scores across cadres and facility types (p>0.05) [Table 4].

**Table 3:** ORIC scores according to change commitment and change efficacy facets.

|  | ORIC scale | Mean score -<br>Pregnancy | Mean score -<br>Postpartum |
| --- | --- | --- | --- |
| **Change**<br>**commitment** | People who work here are committed to implementing IV iron | 4.8 | 4.8 |
|  | People who work here will do whatever it takes to implement IV iron | 4.7 | 4.6 |
|  | People who work here want to implement IV iron | 4.7 | 4.8 |
|  | People who work here are determined to implement IV iron | 4.8 | 4.8 |
|  | People who work here are motivated to implement IV iron | 4.7 | 4.7 |
|  | Mean score (standard deviation)<br>[Mean score%] | 23.7 (2.5)<br>[94.8%] | 23.7 (2.2)<br>[94.8%] |
| **Change efficacy** | People who work here feel confident that the health facility can get people invested in implementing IV iron | 4.7 | 4.6 |
|  | People who work here feel confident that they can keep track of progress in implementing IV iron | 4.8 | 4.7 |
|  | People who work here feel confident that the health facility can support people as they adjust to IV iron | 4.7 | 4.8 |
|  | People who work here feel confident that they can keep the momentum going in implementing IV iron | 4.8 | 4.8 |
|  | People who work here feel confident that they can handle the challenges that might arise in implementing IV iron | 4.7 | 4.6 |
|  | People who work here feel confident that they can coordinate tasks so that implementation of IV iron goes smoothly | 4.7 | 4.8 |
|  | People who work here feel confident that they can manage the politics of implementing IV iron | 4.6 | 4.7 |
|  | Mean score (standard deviation)<br>[Mean score%] | 33 (3.0)<br>[94.3%] | 33 (3.0)<br>[94.3%] |

**Table 4:**
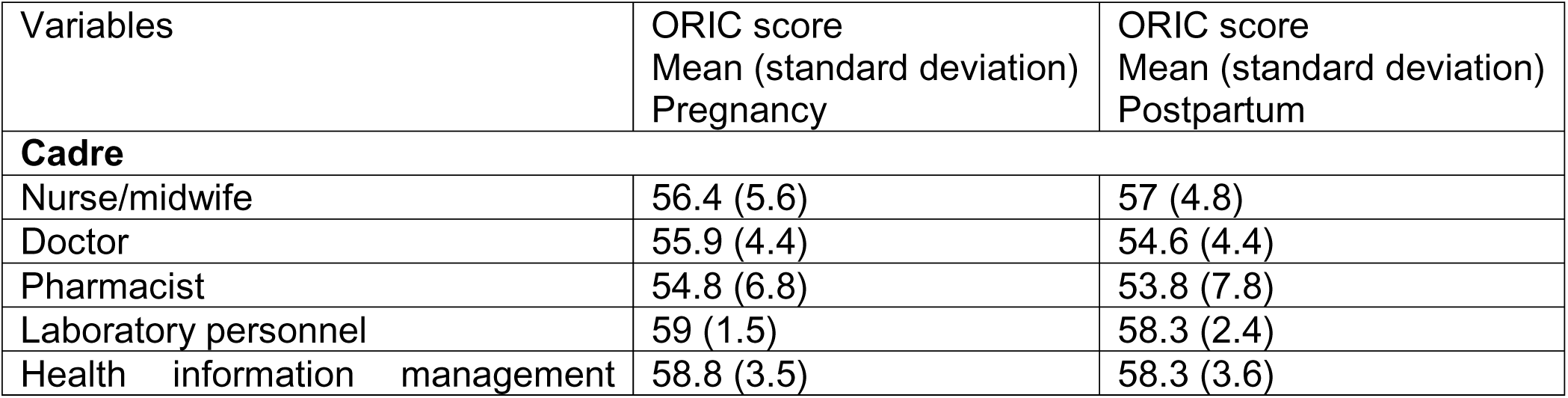
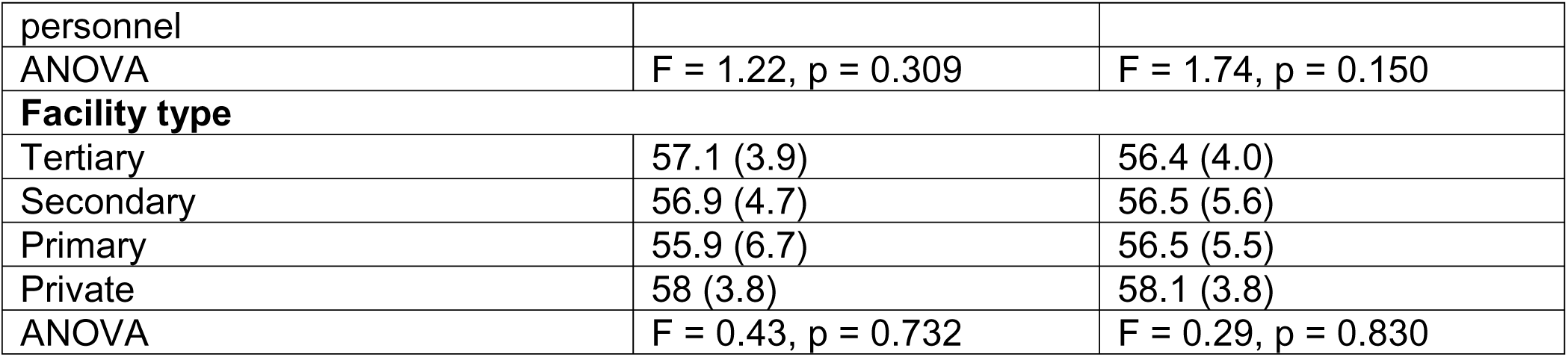
Job cadre, facility type and mean ORIC scores of participants.

### Qualitative results

Sixteen health managers were interviewed, including nine doctors, six nurses/midwives, and a community health worker. Five participants were from secondary facilities, four each from PHC and private facilities, and three from the tertiary facility. Most participants were females (68.8%, 11/16) and within the 50-59 age range (62.5%, 10/16).

Sixty-four HCPs participated in the FGDs, including 37 nurse/midwives and 27 doctors. Thirty-seven were nurse/midwives and 27 were doctors. Five worked at the PHC, 24 at the secondary facilities, 15 at the tertiary facility, and 20 at the private facilities. Most participants were female (71.9%, 46/64) and within the age range of 25 to 39 years (45.3%, 29/64).

Facilitators and barriers are presented according to CFIR constructs and domains and summarised in Table 5.

**Table 5:**
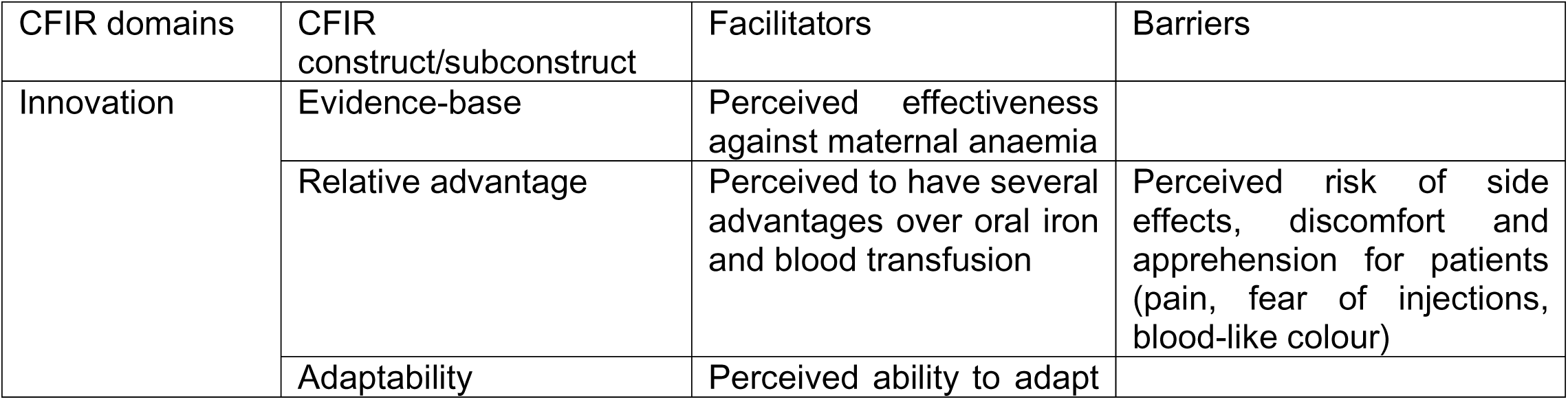
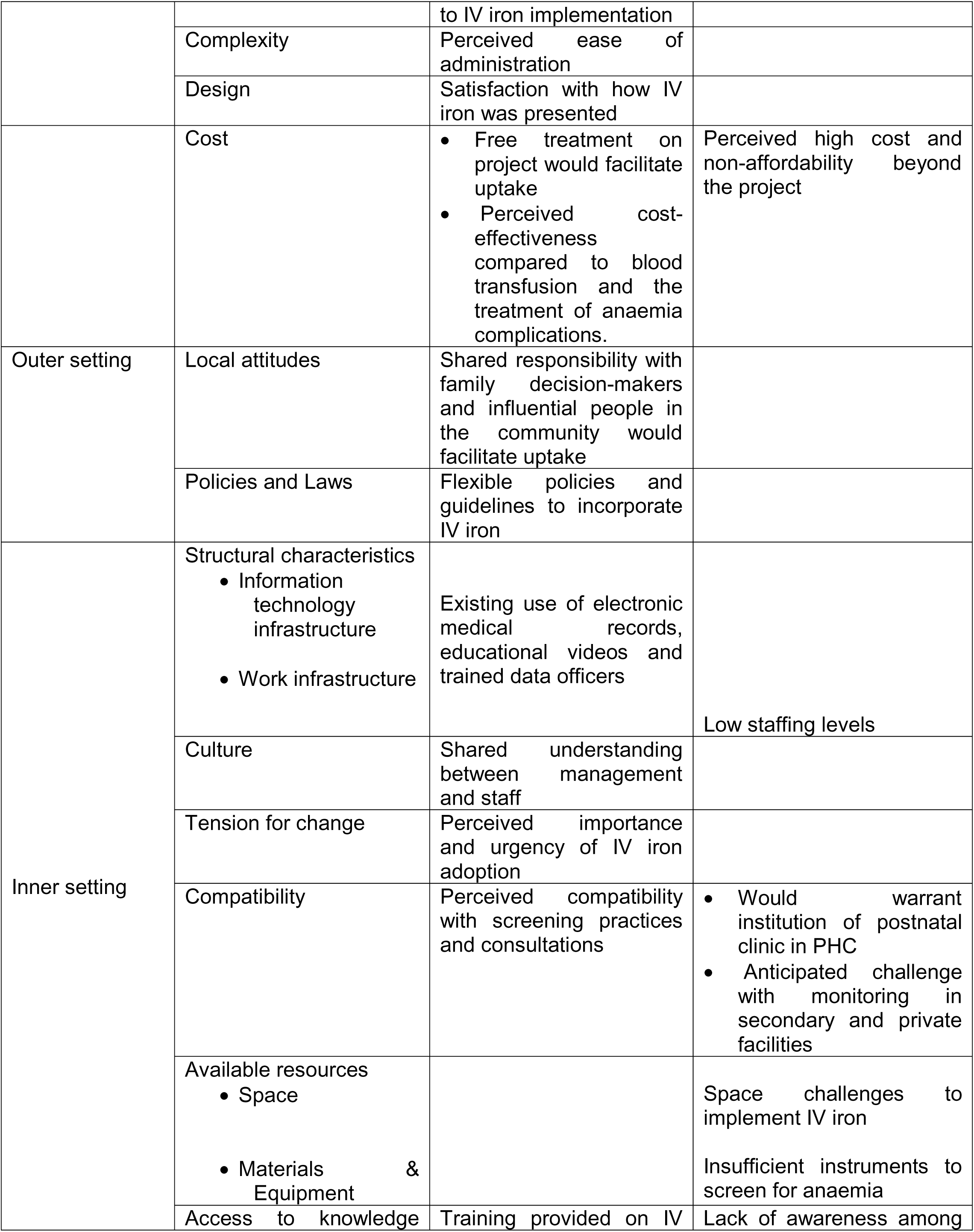
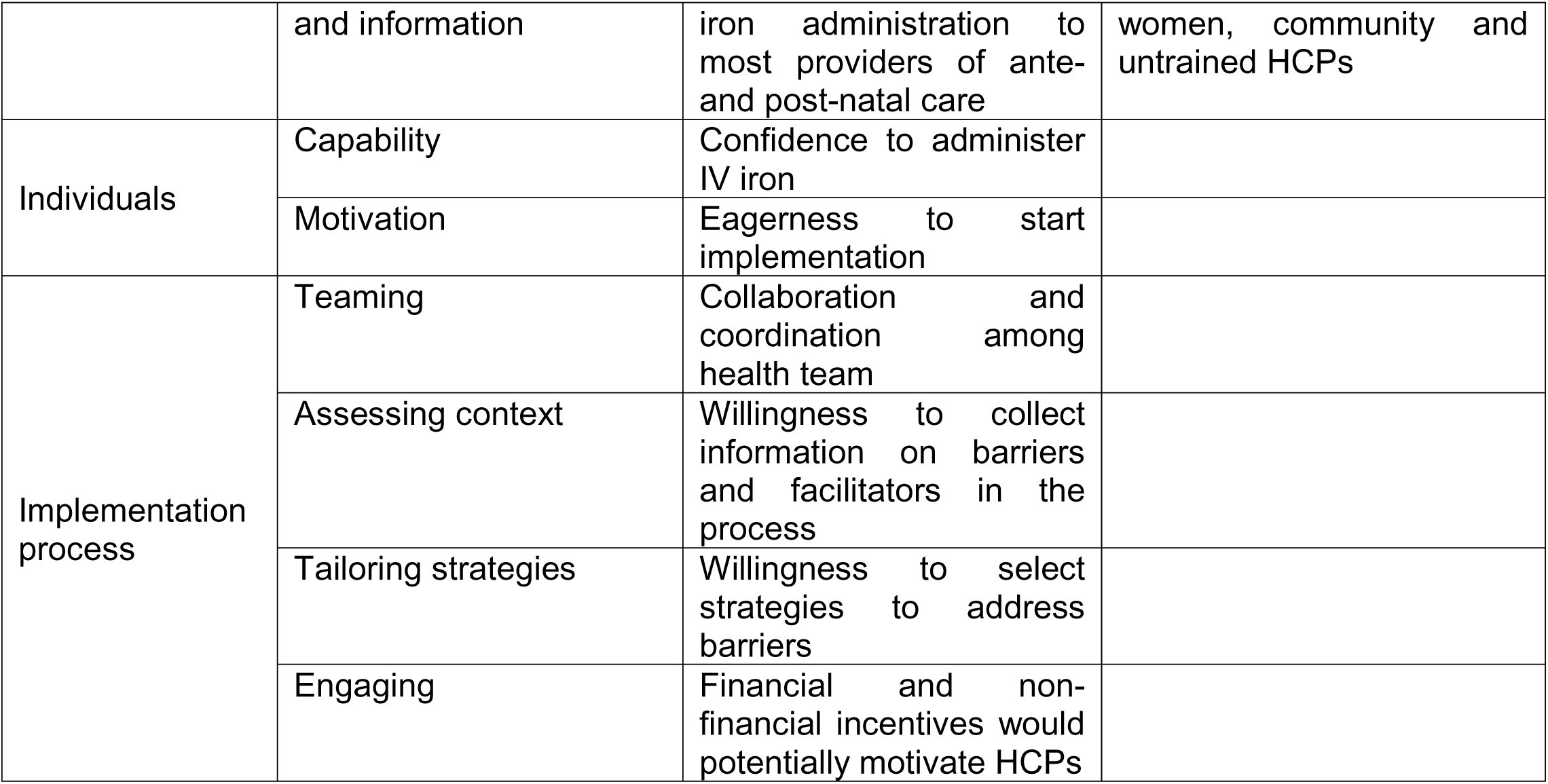
Summary of facilitators and barriers according to CFIR constructs and domains.

### CFIR domain 1: Innovation

#### Facilitators

Construct: Evidence-base. Most of the HCPs perceived IV iron to be effective against maternal anaemia. This was largely due to prior medical training, training on FCM administration and previous use of older formulations. Some HCPs reported on evidence of safety garnered from the training, perceiving that FCM would have fewer side effects than older formulations.

*“Personally, I don’t think there will be much of um, side effects based on what have already been reported. I know that the clinical trial [IVON trial] is different from what we’re doing. What we’re doing now is an implementation study. So, the clinical trial has already answered the question about whether or not the drug is effective and has answered the question about whether or not there’ll be side effects so that we don’t expect much of side effects. We are going to see how that plays out in our own scenario in [Facility X] and we hope that we get the same result*.” [Doctor, male, 25-39 years, secondary facility]

Construct: Relative advantage. Majorly, HCPs perceived that IV iron corrects anaemia faster and more effectively than oral iron, and it addresses the issue of compliance common with oral iron since it is given once and well absorbed. It was also perceived to be a better treatment because of the nausea and vomiting of pregnancy, intolerance to oral iron by some women and the pill burden of oral iron, especially women who are prescribed double-dose haematinics for moderate anaemia. Compared to blood transfusion, IV iron was perceived to be a safer alternative with fewer side effects, cheaper, more acceptable to women particularly those with religious barriers, requires less time for administration and not requiring hospitalisation. The HCPs perceived that IV iron would reduce the need for blood transfusion and associated tests, as blood is not readily available and people are reluctant to donate blood. IV iron is thought to be beneficial to not only the mother in pregnancy, delivery and postpartum, but also to the child, HCWs, community and national development. The HCPs at the PHC said it will promote the image of the facility and reduce the need for referrals to secondary facilities, which is the usual practice for moderate to severe anaemia. Some HCPs in the PHC and private hospital perceived that IV iron would attract more patients to the facility.

*“The benefit is, there’s a benefit to the patient, the doctor and also to the society too… The measure of development is, you check erm (pauses) maternal and peri-natal morbidity and mortality is a major criterion for any society. So, anaemia kills, so if we are able to use this iron formulation to correct anaemia (claps hands), doctor will be happy, doctor will be fulfilled. The confidence will be there. Patient will live, and the statistics also improve and the way they see us and see Nigeria, Africa will also change. It will reflect on our status quo. So, the benefits cut across board. Everybody will be happy at the end of the day because the whole thing we want to achieve result. It is result-oriented.”* [Doctor, female, 25-39 years, private facility]

Construct: Adaptability. All HCPs reported their ability to adapt to IV iron implementation. The potential adaptations for optimised screening and early identification of moderate to severe anaemia for FCM treatment included the use of point-of-care tests, and taking screening from laboratories to antenatal (ANC) clinics and postpartum wards. In consideration of time, some HCPs in the secondary facilities suggested giving FCM as a subcutaneous or intramuscular injection. In consideration of side effects, some HCPs in private and tertiary facilities suggested an increase in standard monitoring time post-infusion. Other suggestions included batching patients and giving FCM on specific days, and administration by designated staff.

*“I do not see a challenge with that, because give or take, this [FCM] should be for every patient that requires it. We’re looking at maybe about an hour from the time of the decision to administer the IV iron. What we could potentially do is have specific days where the hospital is lighter, because we do work based on schedule.”* [Medical director, female, 40-49 years, private facility]

Construct: Complexity. Most HCPs reported no potential challenge in implementing FCM because of prior study training, previous use of IV iron (tertiary facility), and adequate preparation for implementation. It was also perceived to be easy to administer because it is given once without the need for hospitalisation and because it is less cumbersome than blood transfusion. The requirements for monitoring patients were not perceived to be tedious by most staff, who reported having to do it with similar intravenous treatment and even longer monitoring with blood transfusion.

*“It reduces all the cumbersome, all the activities you go into when you want to transfuse. It will reduce it. You must get giving set, you must collect hydrocort, you must collect this one, then the blood has to be at the temperature, body temperature before you can transfuse. All those delay that you have to wait and follow is not there.”* [Nurse, female, 50-59 years, tertiary facility]

Construct: Design. Most HCPs were satisfied with how FCM was presented to them except few participants in a private hospital that were not part of the study training. The trained HCPs perceived the training to be well planned, detailed and allowed for practical training on screening for anaemia and administration of FCM. They also appreciated that there was a step-down training at the facilities, which allowed other staff to be trained. Some noted that the presentation could be improved by using real patients for demonstration and having a video recording of the training. There were suggestions of multiple ways in which FCM could be presented to end-users such as the use of leaflets/posters, radio, social media and the use of educational videos during ANC clinics.

*“In fact, well planned, well planned. After the community stakeholder meeting, we were invited to a meeting, full detail of the process of the research. When we were trained, we now came back to the facility to train the IMT [implementation management team] members, after that we are called for another meeting where we were taught on the screening procedure, the criteria, the stocking, the consent form, then after that they now even came again to the facility to assess our constraints. I just like the outreach survey, I like the planning. It’s well-organized, this is the first time I’m really meeting with well-organized researchers.”* [Senior matron, female, 50-59 years, PHC]

Construct: Cost. Some providers said that providing IV iron for free on the project would facilitate its uptake. In a broader sense, there was also the perception among some HCPs that IV iron could be cost-effective, as it would reduce the likelihood of complications of anaemia such as postpartum haemorrhage and subsequent treatment. It would also reduce the need for costs associated with blood transfusions, such as blood tests and hospital admissions.

*“Yes just like I said before these patients that are on blood transfusion now assuming we have these iron intravenous iron than while she was coming to the antenatal clinic it is something we will administer and the patient will be discharged few hours after that there will be no reason for patients to be admitted to spend money on bed and other things do you understand with this it will caution the effect of, it will caution expenses, the money the patient will have spend on admission, if patients comes in with these anemia the treatment can be given on out patients basis so she save her money on hospital admissions.”* [Senior matron, female, 50-59 years, secondary facility]

#### Barriers

Construct: Relative advantage. There were significant concerns about the side effects of FCM being a barrier to adoption despite the evidence provided during the study training. Some HCPs at the private hospitals expressed fear of losing patronage because of side effects. Other concerns about IV iron compared to oral iron were pain, fear of injections, and the colour of IV iron being mistaken for blood. Also, compared to blood transfusion, IV iron was perceived to take longer to correct anaemia. There was a perceived lack of benefit for specific groups of patients such as those with mild anaemia, sickle cell anaemia, symptomatic anaemia and asthma.

*“Well because of a new medication as it were, what may stand out for me particularly being a private hospital is the fall out if any, you know on towards adverse event occur. It could be a recipe for disaster being a private hospital because word you know travels fast. It could potentially lead to loss of patient because they tend to talk to each other. So that could be a potential problem.”* [Medical director, female, 40-49 years, private facility]

Construct: Cost. For IV iron to be used sustainably, a significant barrier mentioned was cost. In the private hospitals, where many patients are on health insurance, there was a concern about cost to the health management organisations, which would influence their inclusion of IV iron in health plans. HCPs suggested strategies to reduce cost such as government subsidy, health insurance, corporate philanthropy, and direct purchasing in bulk from manufacturers.

*“Since that they said it’s free for now. When the organisation removes the project, and we continue, I don’t know how it’s going to be easy for our patient because of the monetary aspect that they are going to do it, because before they are doing their PCV 500 naira abi? (Right?) And now the moment the programme started they are doing it free and people like free things. But the moment that we tell them that is no more free again, they will tell you that I don’t want to do it.”* [Nurse, female, 40-49 years, PHC]

### CFIR domain 2: Outer setting

#### Facilitators

Construct: Local attitudes. According to HCPs at the PHC, engagement with family decision-makers like spouses and mothers-in-law would facilitate uptake by end-users. Feedback to the community by patients with a positive experience would also facilitate general acceptance, and influential people in the community could encourage women to attend ANC and receive IV iron.

*“There are some patients that irrespective, they have some traditional belief that has actually guided them, or some that their mother in-law is the one making decision in their family, and you know these mothers, they are not well informed. So I don’t know if there is someone that can do follow-up, I can’t really go to their house, but if you have someone who can follow-up, go to their house to tell them these are the benefits, or even show them someone who has taken this thing before, so as for the patient to benefit from it.”* [Midwife, female, 25-39 years, PHC]

Construct: Policies and laws. There were differences in views about the existence of policies for anaemia management. For example, the managers at the PHC and tertiary facility said they use the state policy and departmental policy respectively while staff say they do not have specific policies. A manager at a private hospital said there was no specific policy and treatment is from personal knowledge, while the doctors said they use WHO guidelines. Most of those with a policy said IV iron is a good fit for it and would bring improvement. For the tertiary and secondary facilities, the managers said it was already captured in their professional guidelines. IV iron implementation was also thought to be in alignment with international guidelines.

*“Okay, um, I think it [IV iron] is going to facilitate it [policy]. It is going to make things better, you know, the whole, in some situations where we can foresee a possible need for even blood transfusion, we can start with the IV iron in case and keep things running up until then or even in selected patients that don’t take blood So, there is going to just be a…. an improvised one that factors in IV iron at the stage and level of the anaemia depending and correlating with the patient’s symptoms and the patient herself.”* [Doctor, female, 25-39 years, private facility]

### CFIR domain 3: Inner setting

#### Facilitators

Sub-construct: Information technology infrastructure. All but one of the two secondary facilities and the PHC had electronic medical records already in use, where data on IV iron use can be integrated. One private hospital uses educational videos in the reception of ANC clinics, which HCPs said can incorporate IV iron education. At the PHC, there is a trained data officer ready to collect data relevant to IV iron implementation in registers.

*“For instance, we already have an antenatal video for our patients. Whenever they come for antenatal, they watch a video at the reception so you can find a way of chipping in intravenous iron during that video such that they are sensitized already, that should this happen, I need it.”* [Doctor, female, 25-39 years, private facility]

Construct: Culture. Most HCPs said there was shared understanding between management and staff regarding the implementation of IV iron and that both management and staff were receptive to it. However, one HCP in a private hospital pointed out that they would resist IV iron if adequate plans for side effects were not put in place.

*“Okay I said my MD, that’s the medical director of this facility, and manager, they are aware of this program. They are fully involved so I believe they are, of course, they are very receptive for us to be part of this. So they are very receptive then on the other staffs, the other medical staffs we just need to keep training and retraining but at the moment we are all receptive of this.”* [Senior matron, female, 25-39 years, private facility]

Construct: Tension for change. Adopting IV iron was perceived to be important and urgent by most HCPs because of non-compliance to oral iron by patients, the need to boost haemoglobin promptly before delivery, a needed alternative for blood transfusion, and the need for HCPs to use a long-forgotten intervention. In addition, for the PHC, there would be no need for referrals and loss of patients.

*“It’s important, it’s very, very important because number 1, we will not refer our patients again, and the moment we refer them, some of them don’t come back again. So, it will be a continuity of care, and we care for them from antenatal to the postpartum and it’s our, it’s a thing of joy to us to see our patients. I believe it’s very, very good and me particularly, I love it.”* [Nurse, female, 40-49 years, PHC]

Construct: Compability. Most HCPs did not foresee any significant change in routine or negative effect on consultation with IV iron implementation. Some expressed that it could positively affect consultation by either reducing the number of patients with anaemia requiring follow-up care or by increasing the number of patients who attend ANC because they have an alternative to blood transfusion.

Additionally, IV iron was perceived to fit well with screening practices as it can be given immediately a woman is diagnosed. Two managers in a private and secondary facility said it would not take much time to administer compared to blood transfusion and older formulations of IV iron.

*“Well, I’ll say it fits perfectly because we do a routine, so I’ll say it fits perfectly because within our facility we do amm PCV check that’s anaemia check for pregnant women and Immediate postpartum women so it fits perfectly so it’s easy for us to detect any woman that is anaemic during pregnancy and postpartum [and] administer, it’s easy for us to administer the IV Iron at the same time.* [Senior matron, female, 25-39 years, private facility]

Construct: Access to knowledge and information. Most HCPs were trained by the research team on IV administration and awareness was created among patients at the PHC. There was a consensus that more awareness should be created among patients and continuous training among HCPs for successful implementation.

*“That shouldn’t be a problem for the facility, we have been trained and definitely the doctor will be around and the nurse has to be by the patient. We have been trained to know that the whole infusion should run for over 20minutes and then we are to check vital signs to make sure that the patient is not reacting.”* [HOD, male, 50-59 years, secondary facility]

### Barriers

Sub-construct: Work infrastructure. Staffing level was a consistent identified barrier across facilities. The facilities are short-staffed due to the migration of HCPs and high staff turnover. Many HCPs complained of heavy workload and identified the need for continuous training of new staff.

*“I think the biggest hindrance is brain drain. So even after training people who, after you train people on use of the drug, if those people travel abroad, we have to train other people again, I think that’s the major issue that can come up or if doctors are transferred to other centres and are not replaced, you know, that can also be a challenge and there are things we can’t that are beyond our control.”* [Doctor, male, 25-39 years, secondary facility]

Construct: Compability. Some HCPs in private hospitals anticipate that IV iron implementation will require significant change in routine with more manpower, time and equipment to monitor its administration. For the PHC, it would require the institution of a postnatal clinic as they do not routinely have one. HCPs in secondary and private facilities anticipated challenges in monitoring IV iron administration.

*“In case of IV iron for 15 min, it could be a problem for doctors. maybe he wants to leave and you can’t leave the nurse alone because the nurse there is not the one inside the ward there might be nurse that is part of the implementation flow but I don’t know maybe overtime we feel tired or having some kind of unwillingness waiting to monitor 30 mins few mins after the patient except it’s within work hour between 8 and 3pm or 4pm of the day.*” [Doctor, male, 40-49 years, secondary facility]

Sub-construct: Space. The managers said there is enough space for IV iron implementation, while doctors and nurses in private, secondary and tertiary facilities complained of space challenges particularly when the facilities are busy.

*“What we saying now is the nurses in particular and the space because at times at the ANC like 5 or 6 patient that is going to have the IV something [iron]. Where are we going to put the 5 or 6? We are talking about the space.”* [Nurse, female, 50-59 years, secondary facility]

Sub-construct: Materials and equipment. HCPs in the PHC and a secondary facility complained of a lack of enough instruments to screen patients for anaemia. HCPs at the PHC also reported not having enough drip stands and personal protective equipment like gloves and scrubs. Point-of-care testing was identified as critical for optimal screening at the PHC and one private hospital.

*“We still need more, then um, the instruments to be used to determine what’s going to now lead to the use of the drug. So that’s the centrifuge for screening of the patients. So I think those two, I think because I know there are times when our, um, full blood count machine usually has fault and patients have to wait a longer period to get the results. So if the results can be done within 30 minutes, one hour patient gets the results and within three hours patient already knows intervention she is going to have to improve their health.”* [Doctor, female, 25-39 years, secondary facility]

Construct: Access to knowledge and information. Low awareness among patients, communities and untrained staff in other units/departments was a reported barrier to uptake. One doctor in a secondary facility remarked on the low literacy of women in the surrounding community impeding acceptability. There was concern about being ill-prepared for IV iron implementation among a few practitioners from private and secondary facilities who were not trained.

*“But the biggest challenge I foresee is that [Community X], there the literacy level is low and so acceptability of the drug may be an issue um, for those who may not like to receive IV iron or they don’t even, maybe they’ve never heard of it before, you know, “What is this drug we are talking about?” It may take a while to educate those patients. So, getting people who even though they’re eligible, getting people to actually accept the drug may be a challenge.”* [Doctor, male, 25-39 years, secondary facility]

### CFIR domain 4: Individuals

#### Facilitators

Construct: Capability. Most of the HCPs except for the few who had not been trained felt confident in administering IV iron due to their medical/nursing training, prior experience giving similar intravenous treatment, years of work experience and IV iron training received.

*“I’m confident because we have been taught about how to administer it. Everybody will be doing it right because we are used to administration of IV drugs. We are used to administration of, um, transfusion of blood.”* [Nurse, female, 40-49 years, secondary facility]

Construct: Motivation. Most HCPs were ready to implement IV iron and expressed eagerness to start. Some HCPs in primary and secondary facilities complained about the time it was taking for the implementation to start, citing missed opportunities to use IV iron among women with anaemia. A doctor in the tertiary facility said he was eager to use IV iron so he could be among the first people to adopt the innovation.

*“We have a WhatsApp group, we update ourselves, we go round to monitor them [HCPs]. Everybody is aware, everybody is ready. We are ready, we are on top of our toes. We are waiting for IVON-IS [research team] to come and start the implementation.”* [Senior matron, female, 50-59 years, PHC]

### CFIR domain 5: Implementation process

#### Facilitators

Construct: Teaming. Roles for IV iron administration were well defined across all facilities. HCPs reported being well-coordinated, having been prepared by the research team and collaborating with the health team regarding the project. The suggested promoters of collaboration were continual training, effectiveness and safety of IV iron, steady supply of IV iron and consumables, sufficient manpower, good communication, prioritising patients’ needs, mutual respect, less interprofessional rivalry and working without superiority.

*“Okay as the head of nursing department, I collaborate with other health team members I believe we are in good amm communication and relationship about this project, everyone is aware what this is about, and we are ready to start the process.”* [Senior matron, female, 25-39 years, private facility]

Construct: Assessing context. Information to identify facilitators and barriers to IV iron implementation should be collected using interviews, questionnaires, observation, hospital records, and reports from stakeholders, according to the HCPs. Most of them felt the health team should collect information on barriers and facilitators. In contrast, a few managers in private and secondary facilities felt the research team should collect the information.

*“Like I said in collaboration, it will be coming from different departments. The lab own has to report its own, then the nurses, the doctors, and even the person that is collecting the data, the pharmacists where the drug itself is stored. So, it will be a collaborated effort to get the bottleneck.”* [Nurse, female, 40-49 years, private facility]

Construct: Tailoring strategies. The HCPs identified how strategies to address barriers could be selected with most of them saying members of the health team, or more specifically the IMT, should select strategies to address barriers. Though a few felt the hospital leadership (e.g HOD and hospital management) should select and operationalise strategies. Health outcomes were a commonly recommended measure of success of the strategies. One doctor in the tertiary facility mentioned that patient acceptance should be used. He said, *“I think we should also take into consideration the tolerability for the patient, and the patient satisfaction at the end of the day.”* [Doctor, male, 40-49 years, tertiary facility]

Construct. Engaging. Many HCPs said positive outcomes of the study (e.g. drug effectiveness, patients’ satisfaction, no adverse effects), and continuous training would encourage the adoption of IV iron.

*“The encouragement that can be given to them comes from the result of what we have done. So, if there are improvement, we administer today, tomorrow improvement occurs, then we can keep on telling them, educating them in the trainings. This thing is really working well you know, disseminate information more amongst us.”* [Senior matron, female, 50-59 years, secondary facility]

Most of the HCPs identified strategies to motivate providers to adopt IV iron. These included financial incentives, souvenirs, improved work environment, availability of equipment and medical supplies, drug effectiveness, training opportunities including scholarships, recognition for performance, supervision and provision of refreshments (e.g food). Some of the managers said there was no need to motivate HCPs. One health manager felt it was inappropriate to reward HCPs but okay to reward patients and, in so doing, encourage HCPs. He said, *“You can still put some form of reward for the patient. It will be inappropriate to reward the healthcare providers directly you know that’s why, that’s what I am trying to explain. You know but if the healthcare provider sees that you are giving something to their patients, there should be some form of gratification for them too. That oh this patient has [anaemia] and they did this, even if it is to reduce the cost of the IV iron or whatever. So, the healthcare provider definitely will be encouraged to see them [patients]. You can’t reward the healthcare providers, you know that will be wrong and that will be inappropriate.”* [Deputy managing director, male, 40-49 years, secondary facility]

## Discussion

There is evidence for the effectiveness and safety of FCM in the treatment of anaemia in pregnant and postpartum Nigerian women (14,15). We assessed readiness and identified facilitators and barriers to the adoption of FCM in real-world settings as part of pre-implementation activities in Lagos, Nigeria. This novel study provides important results to inform implementation strategies for the adoption of FCM to treat maternal anaemia. The results of the ORIC survey showed high readiness across both change commitment and change efficacy facets for IV iron use in both pregnancy and the postpartum period. ORIC scores did not differ significantly across cadre and facility type. The qualitative results revealed several facilitators across all CFIR domains and barriers in the intervention characteristics and inner setting domains.

Organisational readiness is critical for the successful implementation of an EBI (19). High readiness by HCPs to implement IV iron treatment for maternal anaemia implies they would invest more in the change effort, specifically by initiating change, supporting the change process and by persisting in the presence of barriers (20). Similarly, there was high readiness among HCPs to implement a new midwifery model of care in rural South Australia (25). The level of readiness could have been influenced by the facilitators identified in this study.

In the “intervention characteristics” domain, we identified several facilitators such as perceived effectiveness, relative advantage over existing options, adaptability and non-complexity. Providers in previous studies similarly noted the advantages of IV iron over oral iron and blood transfusion (12). HCPs in this study were especially optimistic that IV iron could be a suitable alternative to blood transfusion, which is unacceptable and not readily available or safe for many women in LMICs (31). This view was also shared HCPs in a previous study in Nigeria (16). Newer formulations of IV iron like FCM with shorter infusion times are less complex and could potentially be administered with higher fidelity (32,33). However, the fidelity of FCM administration in controlled trial settings in Nigeria was “moderate” (18), and worth considering for assessment during its implementation in real-world settings. Unlike our findings, HCPs from previous studies in high-income countries reported difficulty with IV iron administration (34,35). The difference may be because these facilities were already implementing IV iron and were sharing actual experiences.

The “inner setting” domain, which is closely intertwined with organisational readiness, also had several facilitators. Organisational culture can influence organisational readiness and implementation success of innovations (36,37). There was a shared understanding about IV iron implementation among management and staff in this study, and HCPs valued IV iron as important for treating maternal anaemia. These are aspects of change valence, which influence change commitment (20). Previous training on IV iron and its compatibility with routine processes would help the HCPs to know what to do and how to do it, which influence change efficacy (20).

The barriers identified in this study were within the “Intervention characteristics” and “inner setting” domains. Some of them were also mentioned in a systematic review, namely, perceived high cost, risk of side effects, patients’ lack of knowledge, fear and apprehension (12). Interestingly, the barriers related to work infrastructure and resource availability did not impact ORIC scores, possibly because there were also several facilitators within the inner setting and the HCPs were highly motivated. Indeed, motivation is considered an important complement to organisational capacity and innovation-specific capacity when making a practical judgement of organisational readiness (38).

While there was no significant variability across facility types for ORIC scores, some facilitators and barriers identified in this study were unique to facilities. For example, family and community engagement was deemed a potential facilitator at the PHC, buttressing previous findings of partner support and community involvement determining the success of anaemia reduction strategies in LMICs (39). PHCs also tend to have fewer resources and a smaller scope of services than higher levels of care as evident in this study where insufficient materials for IV iron implementation and less compatibility with postnatal care services were potential barriers.

Given high readiness scores coexisting with structural barriers and influence of community decision-makers, our study findings warrant a conceptual extension ORIC and CFIR that is tailored for LMIC maternal health innovations (Figure 1).

**Figure 1:**
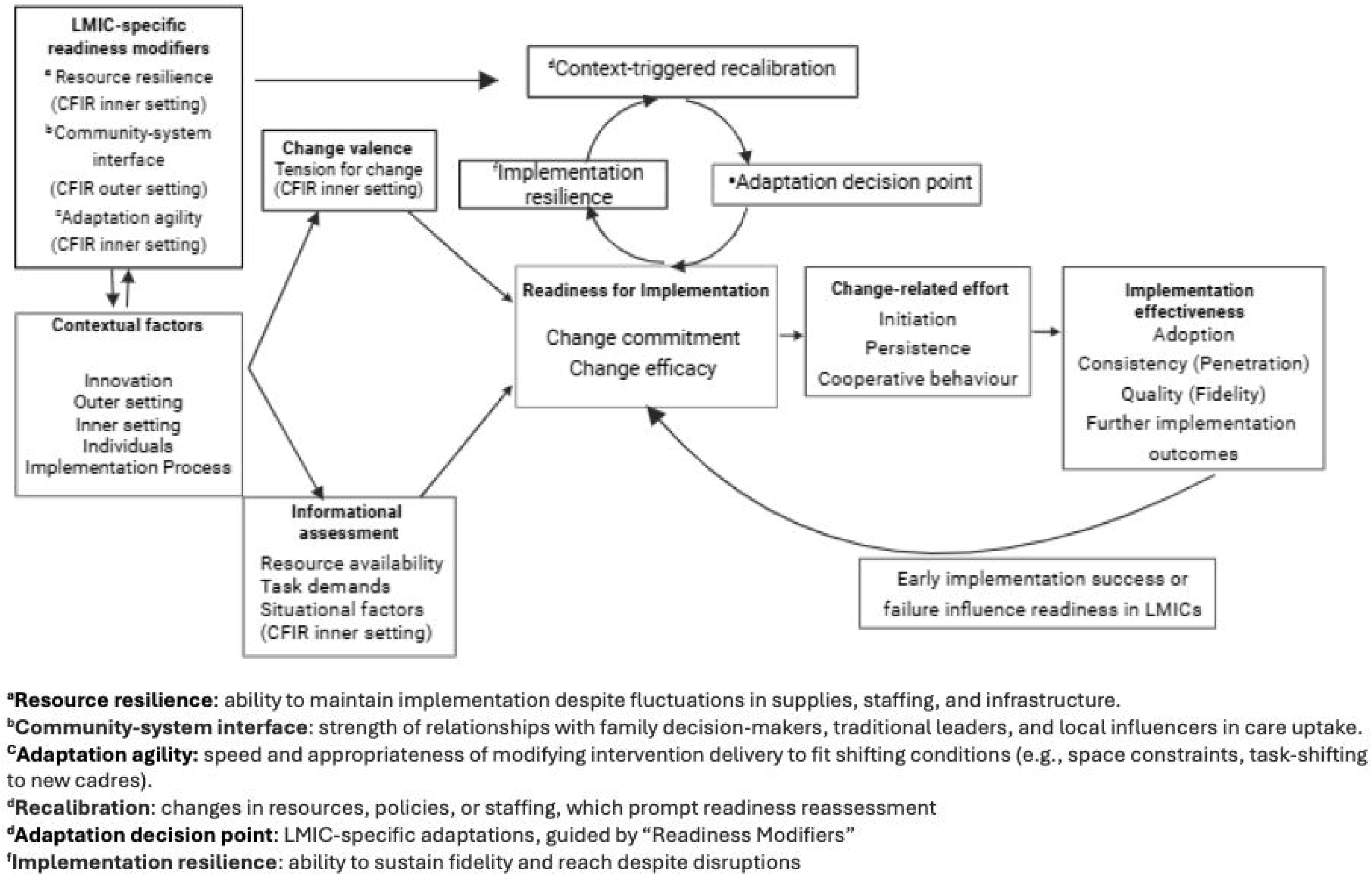
Conceptual model showing interplay of LMIC-specific readiness modifiers with ORIC-CFIR framework (adapted from the Theory of Organisational Readiness for Change-CFIR integrated framework(19,40))

Our adapted model suggests that readiness modifiers such as resource resilience, community-system interface and adaptation agility in LMIC settings are linked to CFIR domains and can trigger recalibration, adaptation and implementation resilience despite barriers. These would readiness for implementation in LMICs. Similarly, early implementation success or failure after these efforts, would influence readiness in LMICs and determine if change-related effort is sustained. We suggest that this adapted model could guide pre-implementation assessment, adaptive strategy selection, and monitoring of readiness over time in LMICs settings.

### Implications for practice, policy and future research

Given the readiness and facilitators identified in this study, there is a high potential for IV iron to be adopted for the treatment of maternal anaemia in the study sites. However, for successful implementation, tailored strategies are required to address identified barriers. For example, the perceived risk of side effects despite the overwhelming evidence of the safety of IV iron stems from prejudice against older formulation that had high rates of adverse events(41). This can be addressed by continuous training of HCPs, which could also address the issue of high staff turnover. Training helps onboard new staff who replace previously trained staff but is also effective as a retention strategy for HCPs(42). Education of patients is also critical, leveraging on technology such as videos where available. Community education through social and mass media is equally important.

To address space challenges in facilities, particularly when multiple women require the treatment at the same time, existing spaces can be optimised bearing in mind the needs of both patients and staff and working with experts in healthcare design and space management for maximal benefit (43). Adaptability of IV iron, while feasible, should be based on the context of the healthcare facilities, considering specific resources and characteristics. Adaptation should also be ongoing through the implementation process due to dynamic settings and needs. The use of an implementation framework to systematically inform the process is also recommended (44,45).

For successful scale-up, cost reduction strategies are required for IV iron to be used sustainably beyond funded projects. Suggested strategies by HCPs such as government subsidy and health insurance should be considered (16). For universal access, another important consideration is the inclusion of IV iron in the national essential medicines list by policymakers. This will facilitate government financing to ensure its inclusion in benefit packages in the public sector and health insurance schemes (46). Donors and funders are also vital in providing financial and technical support and facilitating collaboration across the health system. In addition, the inclusion of IV iron in maternal anaemia management guidelines by health systems planners and professional associations would support safe and standardised use of IV iron.

The suggestions by HCPs in this study to motivate them to implement IV iron resonated with the themes identified by Willis-Shattuck et al as motivating factors in LMICs. These included financial rewards, career development, continuing education, hospital infrastructure, resource availability, and recognition/appreciation(47). While use of financial incentives may be successful in motivating HCPs (48), it is not sustainable as a strategy to implement every innovation. Non-financial incentives should be explored in implementing IV iron as these are realistic and have a greater chance of integration into local budgets (49). Lessons can be taken from non-financial incentives that work in LMICs. For example, leadership and supervision, continuing education, and availability of infrastructure and resources were significant predictors of motivation for HCPs in Ghana (48)

Future research should evaluate and address barriers in the implementation phase of IV iron, potentially using CQI strategies (23), which foster continuous learning, innovation and improvement (50). However, these CQI strategies require necessary structure, resources, and resolve for effective execution (51), all important considerations for the IVON-IS project.

There is evidence that multiple-dose IV sucrose is more cost-effective than oral iron therapy for maternal anaemia in LMICs (52). However, similar research using FCM is limited but necessary to provide convincing evidence to policymakers, providers, and end-users to adopt it for the treatment of maternal anaemia.

### Strengths and limitations

This is one of the few studies to assess organisational readiness, facilitators and barriers to IV iron adoption in real-world settings in LMICs. We used a mixed-methods design to collect data from HCPs who would directly be involved in the implementation of IV iron, and we got a fairly high response rate from eligible participants in our online survey. Using CFIR to guide our data collection and analysis, including adapting to the updated CFIR and using inductive-deductive coding approaches, strengthened our reporting of facilitators and barriers.

However, our findings cannot be generalised to facilities across Nigeria. Lagos state is more cosmopolitan than other states in the country, where the determinants of adoption of IV iron may vary. Also, there is the potential for selection bias in the online survey as those who chose to participate may differ from those who did not, for example, in their motivation. There is also the possibility of social desirability bias in the provision of favourable responses. In collecting qualitative data, we only evaluated input from doctors and nurses. We omitted other cadres that had participated in the survey such as pharmacists and laboratory personnel, who may have had useful insights into the topic.

## Conclusion

This research affirms the readiness of Nigerian health systems to adopt IV iron for maternal anaemia, provided that identified barriers are systematically addressed. Emphasising provider and patient education, cost reduction strategies, continuous quality improvement, and contextual adaptation will be key to realising the potential impact of FCM on maternal health outcomes in Nigeria.

## Data Availability

All data produced in the present study are available upon reasonable request to the authors

## Acknowledgements

We acknowledge Dr. Ejemai Eboreime for his contribution towards the design for this study and Damilola Onietan for her administrative role in supporting field activities. We appreciate the research assistants, Bisola Omotayo and Prince Godson, who supported qualitative data collection. Finally, we are grateful to the health care providers who took time to share their perspectives in this study.

